# Investigating the relationship between IGF-I, -II and IGFBP-3 concentrations and later-life cognition and brain volume

**DOI:** 10.1101/2020.10.09.20209833

**Authors:** Antoine Salzmann, Sarah-Naomi James, Dylan M. Williams, Marcus Richards, Dorina Cadar, Jonathan M. Schott, William Coath, Carole H. Sudre, Nishi Chaturvedi, Victoria Garfield

## Abstract

**Background:** The insulin/insulin-like signalling (IIS) pathways, including Insulin-like Growth Factors (IGFs), varies with age. However, their association with late-life cognition and neuroimaging parameters is not well characterised.

**Methods:** Using data from the British 1946 birth cohort we investigated associations of IGF-I, -II and IGFBP-3 (measured at 53 and 60-64 years) with cognitive performance (word learning test (WLT) and visual letter search (VLS) – at 60-64y and 69y) and cognitive state (Addenbrooke’s Cognitive Exam-III (ACE-III) - at 69-71y), and in a proportion, quantified neuroimaging measures (whole brain volume (WBV); white matter hyperintensity volume (WMHV); hippocampal volume (HV)). Regression models included adjustments for demographic, lifestyle and health factors.

**Results:** Higher IGF-I and IGF-II at 53y was associated with higher ACE-III scores (ß 0.07 95%CI [0.02,0.12]; score_ACE-III_ 89.48 [88.86,90.1], respectively). IGF-II at age 53y was additionally associated with higher WLT scores (score_WLT_ 20 [19.35,20.65]). IGFBP-3 at 60-64y was associated with favourable VLS score at 60-64y and 69y (ß 0.07 [0.01,0.12]; ß 0.07 [0.02,0.12], respectively), higher memory and cognitive state at 69y (ß 0.07 [0.01,0.12]; ß 0.07 [0.01,0.13], respectively) and reduced WMHV (ß -0.1, [-0.21,-0.00]). IGF-I/IGFBP-3 at 60-64y was associated with lower VLS scores at 69y (ß -0.08, [-0.15,-0.02]).

**Conclusions:** Increased measure in IIS parameters (IGF-I, -II and IGFBP-3) relate to better cognitive state in later life. There were apparent associations with specific cognitive domains (IGF-II relating to memory; IGFBP-3 to memory, processing speed and WMHV; and IGF-I/IGFBP-3 molar ratio with slower processing speed). IGFs and IGFBP-3 are associated with favourable cognitive function outcomes.

## Introduction

The Growth Hormone/Insulin-like Growth Factor signalling pathway is thought to play a key role in ageing health and the determination of age-related diseases. In humans, this pathway is regulated through the somatotropic axis, which consists mainly of growth hormone (GH), Insulin-like Growth Factors (IGF) and their respective binding proteins ^1^. IGF-I binds to IGF binding proteins (IGFBP) in high-affinity complexes to prevent degradation and binding to the IGF-I receptor (IGF1R), thus leaving concentrations of ‘free’ circulating IGF-I relatively low. As such, the ratio of IGF-I to binding proteins provides an indication of bioavailability ^1–3^, whilst also mediating its effect via IGF1R; IGF-II bioavailability is also regulated by the IGF2 receptor ^4^. IGFBP-3, however, has also been shown to have IGF-independent associations on phenotypes ^5^.

Whereas research supports an important role of Insulin/Insulin Growth Factor-like Signalling (IIS) in early brain development and neuroplasticity, as evidenced by developmental defects associated with mutations in the IGF1R gene, its role in the maintenance of brain function is less well understood ^6,7^. Observational studies are conflicting, with some reporting protective, detrimental, ‘U’ shape and null associations ^8,9^. The relationship between IGF-II and cognition and neurological health, on the other hand, remains largely uncharacterised, with only one study to date, which reported raised IGF-II concentrations in association with better cognition, while no association of IGF-I was observed ^10^

The MRC National Survey for Health and Development (NSHD; also known as the 1946 British Birth Cohort), provides an ideal opportunity to study the relationship between measures of the IIS axis through mid-life, and cognition and brain volume in later life ^11^. IGF-I and -II collection at multiple time points may help resolve previous discrepant findings by considering differential associations of age, and duration of follow-up. Additionally the NSHD has a wealth of data across the life-course, allowing us to control for potential confounders which have been omitted or based on recalled data in past studies, such as early-life cognition and affective symptoms. Finally, neuroimaging data enables associations with brain volume and neurovascular health to be studied in tandem with cognitive function.

We aimed to examine the independent associations of IGF-I, IGFBP-3, and IGF-II measured at ages 53 and 60-64 with cognitive function at ages 60-64 and 69, and volumetric and cerebrovascular neuroimaging parameters (total brain, hippocampal and white matter hyperintensity volumes) at age 69-71.

## Methods

### Study participants

The NSHD recruited 5,362 participants born within one week in March 1946 in England, Scotland and Wales. The cohort has remained in continuous follow-up from birth to date, with 24 sweeps of investigation ^12^. Cognitive function at age 69 was our primary outcome. At this age, 2149 participants were interviewed. The loss to follow-up was due to death (19.1%), refusal (20.6%), emigration or being temporarily abroad (10.8%) or because the participant could not be contacted or traced (7.8%). Figure 1 outlines the number of participants with information concerning exposure and outcome variables and the final sample (n=1762) used for analysis.

**Figure 1:**
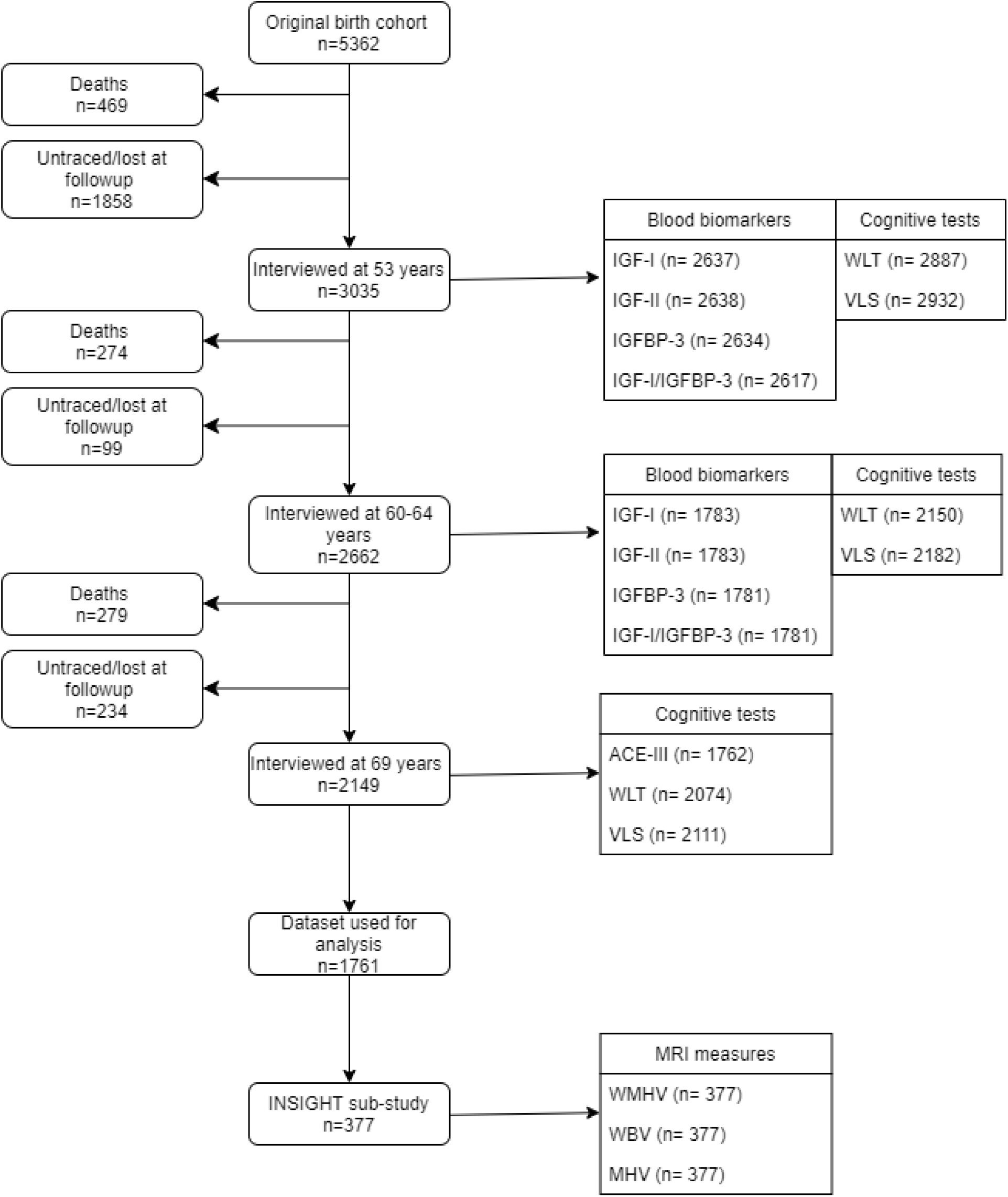
NSHD study flow diagram. Abbreviations: Insulin-Growth Factor-I (IGF-I), Insulin Growth Factor-II (IGF-II), Insulin-Growth Factor Binding Protein-3 (IGFBP-3), Addenbrookes Cognitive Exam-III (ACE-III), Word Learning Test (WLT), VLS (Visual Letter Search) test, Whole Brain Volume (WBV), White Matter Hyperintensity volume (WMHV), Mean Hippocampal Volume (MHV).

We additionally examined associations with neuroimaging outcomes in a sub-sample who had participated in the neuroimaging sub-study, Insight 46 ^13^. Details have previously been published ^13^, but in brief this study included 502 individuals who underwent positron emission tomography– magnetic resonance imaging (PET-MRI) scanning using a single Biograph mMR 3T PET-MRI scanner (Siemens Healthcare, Erlangen), with simultaneous acquisition of PET/MRI data, including volumetric (1.1 mm isotropic) T1 and fluid-attenuated inversion recovery (FLAIR) sequences ^13^. Volumetric T1-weighted and FLAIR images underwent visual quality control prior to processing following an automated pipeline ^13^. Relevant imaging measures were isolated as follows: total brain volume (TBV) using Multi-Atlas Propagation and Segmentation ^14^, mean hippocampal volume (HV) using Similarity and Truth Estimation for Propagated Segmentations ^15^, total intracranial volume (TIV) using SPM12 (Wellcome Centre for Human Neuroimaging) and white matter hyperintensity volume (WMHV) using Bayesian model selection (BaMoS) ^16,17^. WMHV was isolated from 3D T1 and FLAIR images followed by visual inspection to generate a global WMHV, including subcortical grey matter but excluding infratentorial regions ^17,18^. Eligibility criteria, the study protocol, and a comparison of those who did and who did not participate in Insight 46 have been described in detail elsewhere ^13,18^. Of the 502 Insight 46 participants, complete neuroimaging and ACE-III data were available for 378.

## Measures

### IGF-I, IGF-II and IGFBP-3 concentrations

Blood samples were collected at age 53 (non-fasting), and age 60-64 (fasting). Samples were stored at -80°C and assayed together. IGF-I, IGF-II and IGFBP-3 concentrations were obtained by radioimmunoassay using standard protocols in the same laboratory, as previously described ^19^. The intra-assay coefficient of variation (CV) for IGF-I, IGF-II and IGFBP-3 were 3.4%, 2.8% and 3.9%, respectively. The inter-assay CV was 13.7%, 7.4% and 11.7% for IGF-I, IGF-II and IGFBP-3, respectively ^20^. IGF-I, IGF-II and IGFBP-3 values were converted from ng/ml to standard SI units: nmol/L ^21^. IGF-I/IGFBP-3 values were calculated as molar ratios as an indicator of IGF-I bioavailability. Repeat measures were available for between 1117 and 1127 participants, for IGF-I/IGFBP-3 ratio and IGFBP-3, respectively.

### Cognitive performance measures

Three separate tests were used to measure cognitive domains. At 60-64 and 69, standardised versions of a Word Learning Test (WLT) and Visual Letter Search test (VLS) were used to assess memory and processing speed, respectively. The WLT consists of a 15-word learning task devised for the NSHD. Each word was shown to the participant for 2 seconds and partcipants were asked to immediately write down as many words as they could remember. The task was repeated three times, and the scores were summed (max = 45), which is representative of short-term episodic memory ^22,23^. The VLS required participants to scan for the letters ‘P’ and ‘W’ among other letters and cross out as many of them within one minute. Search speed score was based on how far the participant got in the grid search, with a maximum score of 600 signifying they had completed the whole grid ^22^.

The Addenbrooke’s Cognitive Examination-III (ACE-III) is a test for cognitive state administered at age 69 ^24,25^. The test assesses five domains, each with different score ranges: Attention (0-18), Memory (0-26), Fluency (0-14), Language (0-26) and Visuospatial (0-16). The test was administered either online using an iPad (http://www.acemobile.org/) or by paper if the former was not possible. Paper tests were scored by trained personnel ^24,26^. Unlike most tests of cognitive state, the ACE-III total score (max = 100) has a quasi-normal distribution, making it appropriate for parametric statistical analyses.

### Covariates

Covariates were selected *a priori*, based on previous studies and added into our models successively. Model 1 had sex, Model 2 included additional adjustments for childhood cognition and educational attainment, Model 3 added smoking status at the time of exposure and Model 4 added affective symptoms at the time of outcome ^10,27,28^. The same models were used when investigating the relationship between exposures and brain volume with TIV additionally controlled for in all volumetric outcome models.

Childhood cognition at age 8 was assessed by four tests administered by trained observers: reading comprehension, word pronunciation, vocabulary and non-verbal reasoning. Total scores were standardised to the study population. Lifetime education attainment was based on the highest educational qualification by the age of 43 years ^26^ and categorised as 1) no qualification or vocational qualifications 2) ordinary, advanced secondary and 3) tertiary qualifications. Smoking status was obtained from questionnaires and classified into current smoker, ex-smoker and never smoker at age 53 and 60-64 ^29^. Affective symptoms were self-reported at ages 60-64 and 69 years using the 28-item General Health Questionnaire (GHQ-28) ^30^. A threshold, set at the recommended 4 (non-case) / 5 (case) cut was imposed on the GHQ-28 total score to determine case-level symptoms ^30,31^.

### Statistical analyses

WMHV was log_e_-transformed to account for skewness. ANOVA tests were used to examine exposure and outcome variable differences at different timepoints. Within the selected sample, missing data ranged from 0.1% to 27.9% for affective case symptoms (age 69) and IGF-I/IGFBP-3 molar ratio (age 60-64) respectively. To account for attrition at follow-up, missing data for exposure, WLT, VLS and covariate variables were imputed using multiple imputation by chained equations (MICE). Imputation models included all model variables, except for volumetric outcomes. Datasets were imputed 20 times using a Bayesian linear regression model ^32^.

Linear regression models were used to assess the relationship between exposure and outcomes. Results are presented as standardised beta values with 95% confidence intervals. To determine whether the associations of IGFs and IGFBP-3 with cognition and brain volume differed by sex, which may warrant stratification, an interaction term for sex was tested at the 10% significance level for all exposures. No interaction was found between sex and exposure variables, so main models were not stratified by sex. In addition, a quadratic term for the exposure was tested to explore the possibility of non-linear associations. A quadratic relationship was observed between IGF-II and general cognitive state and visual memory. Therefore, for associations with cognitive state, IGF-II measurements were categorised into tertiles.

All statistical analyses were performed in R version 3.5.2 ^33^. Multiple imputation was performed using the ‘mice’ package in R^32^. Relevant R script is available upon request.

## Results

### Sample characteristics

Of the 1762 NSHD participants with cognitive state measured at age 69, 849 (48%) were male, and 913 (52%) were female. The Insight 46 imaging sub-study at age 69-71 (mean age: 70.67 years, SD: 0.67) included 378 of these 1762 NSHD participants. Of these, 194 (51%) were male and 184 (49%) female (Table 1). Whilst those who attended the clinic at ages 53 and 60-64 were overall healthier – less likely to smoke, higher Socioeconomic status (SES) and were more educated –, the concentration of IGF-pathway components was similar between participants in the full sample and those included in the final analysis (data not shown).

**Table 1:** Table presents variable information for the sample (n = 1762 (NSHD) and n= 378 (INSIGHT)) prior to imputation.

|  | NSHD (n=1762) |  |  | Insight 46 (n=378) | P-value<sup>1</sup> |
| --- | --- | --- | --- | --- | --- |
|  | 53 Years | 60-64 Years | 69 Years | 69 – 71 years |  |
| **Exposures** |  |  |  |  |  |
| IGF-I levels (nmol/L) | 26.22<br>(9.00) | 23.00<br>(7.71) |  |  | < 0.001 |
| IGFBP-3 levels (nmol/L) | 173.22<br>(39.31) | 120.11<br>(30.16) |  |  | < 0.001 |
| IGF-I/IGFBP-3 molar ratio | 0.16<br>(0.06) | 0.20<br>(0.06) |  |  | < 0.001 |
| IGF-II levels (nmol/L) | 99.82<br>(32.36) | 87.86<br>(38.60) |  |  | < 0.001 |
| **Cognitive tests** |  |  |  |  |  |
| Addenbrooke's Cognitive Examination<br>(ACE-III) score |  |  | 91.52<br>(6.01) |  |  |
| Visual Letter Search (VLS) score | 283.52<br>(73.34) | 268.25<br>(70.54) | 263.56<br>(74.32) |  | < 0.001 |
| Word Learning Test (WLT) score | 24.87<br>(6.12) | 24.68<br>(6.06) | 22.38<br>(6.13) |  | < 0.001 |
| **Insight 46 MRI measures** |  |  |  |  |  |
| Age at scan |  |  |  | 70.67<br>(0.67) |  |
| Hippocampal volume (ml) |  |  |  | 3.13<br>(0.33) |  |
| Total intracranial volume (ml) |  |  |  | 1432.86<br>(127.58) |  |
| White matter hyperintensity volume<sup>2</sup> (ml) |  |  |  | 3.08<br>(5.19) |  |
| Whole brain volume (ml) |  |  |  | 1101.38<br>(95.34) |  |
| **Model covariates** |  |  |  |  |  |
| Early life cognition | 0.14<br>(0.69) |  |  |  |  |
| **Affective Symptoms Case Status** |  |  |  |  |  |
| Non-case | 1313<br>(81%) | 1247<br>(83%) | 1505<br>(86%) |  |  |
| Case | 316<br>(19%) | 264<br>(17%) | 251<br>(14%) |  |  |
| **Educational attainment** |  |  |  |  |  |
| No qualifications/vocational only | 637<br>(36%) |  |  |  |  |
| O-Level/A-Level or equivalent | 835<br>(48%) |  |  |  |  |
| Higher | 272<br>(16%) |  |  |  |  |
| **Smoking status** |  |  |  |  |  |
| Current smoker | 315<br>(19%) | 148<br>(10%) | 525<br>(9%) |  |  |
| Ex smoker | 842<br>(51%) | 875<br>(58%) | 1061<br>(61%) |  |  |
| Never smoker | 507<br>(30%) | 489<br>(32%) | 525<br>(30%) |  |  |

<sup>1</sup>P-values were calculated using a one-way ANOVA for continuous variables. <sup>2</sup>Median and interquartile range presented for non-normally distributed variables

In the entire cohort at age 60-64, the overall pattern shows that IGF-I concentrations were lower, and IGFBP3 levels substantially lower than at age 53 (Table 1). As a result, IGF-I/IGFBP3 ratio was higher at age 60-64 than 53. IGF-II concentrations were lower at age 60-64 than age 53. Visual memory (WLT) and processing speed (VLS) were lower with increasing age at investigation, as previously described (Table 1) ^34^.

### IGF-I

Higher IGF-I concentrations at age 53 and 60-64 were associated with better cognitive function for the WLT and ACE-III cognitive measures, but not for the VLS (Figure 2). These associations reached conventional levels of statistical significance for IGF-I at age 53 and ACE-III at age 69 (ß 0.07, 95% CI 0.02,0.12, p = 0.008). Adjustment for childhood cognition and educational attainment attenuated this estimate to ß 0.04, 95% CI 0.00,0.09, p = 0.063. Further adjustment for smoking status and emotional symptoms did not alter the estimate (Figure 2). Excluding participants with case-level affective symptoms at baseline had no qualitative impact on the magnitude or direction of associations (data not shown).

**Figure 2:**
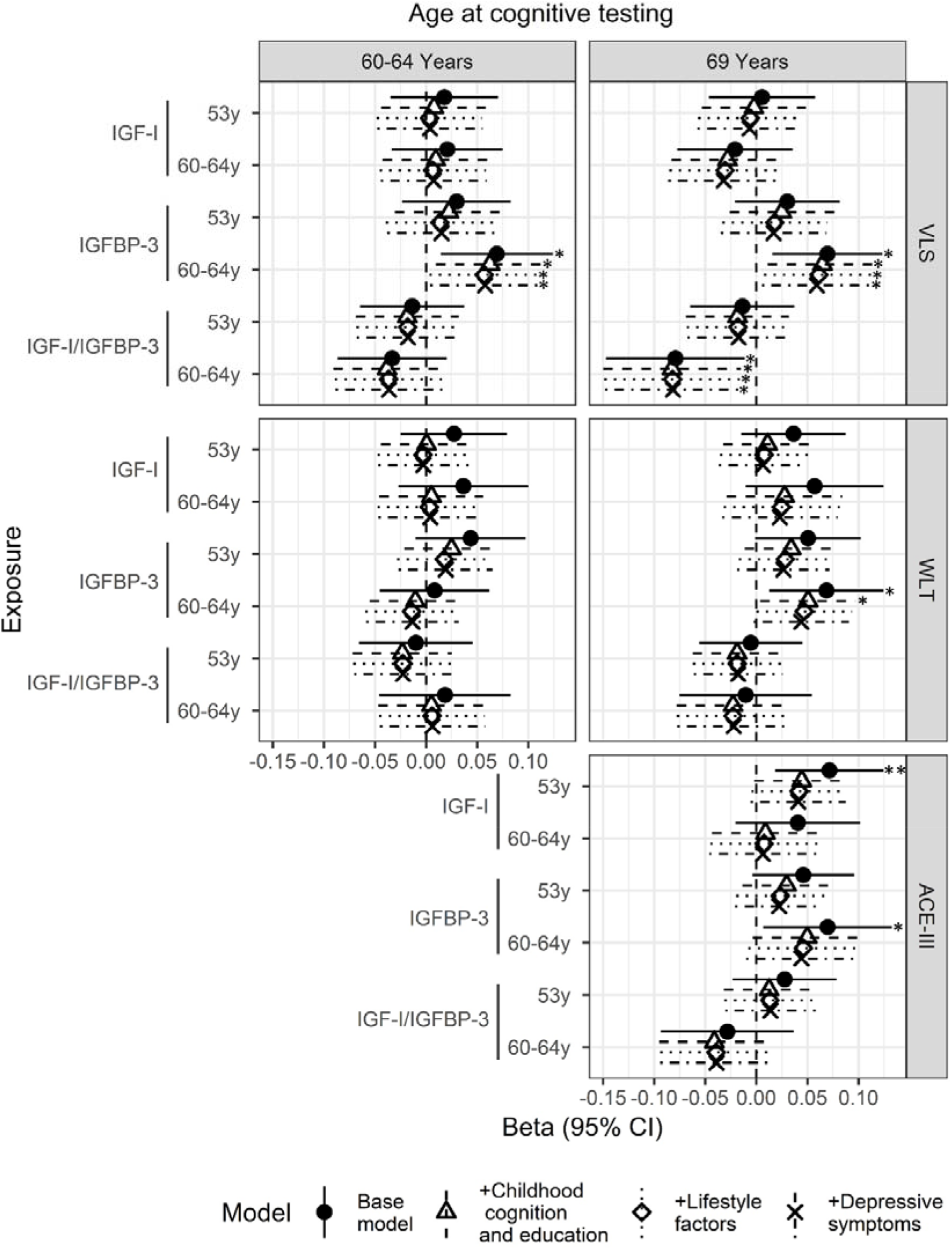
Beta coefficient and 95% confidence interval for the effect of IGF-I, IGFBP-3 and IGF-I/IGFBP-3 at ages 53y and 60-64y on visual memory (WLT) and processing speed (VLS) at ages 60-64y and 69y and cognitive state (ACE-III) at age 69y. Models were adjusted for sex (base model), sex, early life cognition and lifetime education (+ childhood cognition and education model), sex, early life cognition, lifetime education and smoking (+ lifestyle factors) and sex, early life cognition, lifetime education, smoking and depressive symptoms (+depressive symptoms). Abbreviations: Insulin-Growth Factor-I (IGF-I), Insulin-Growth Factor Binding Protein-3 (IGFBP-3), Addenbrookes Cognitive Exam-III (ACE-III), Word Learning Test (WLT), VLS (Visual Letter Search) test. * significant at p < .05, ** significant at p < .01.

In the subjects with imaging available, higher concentrations of IGF-I at age 60-64 were directionally associated with a 8% lower WMHV at age 71 years (95% CI -19%, 3%, p = 0.142), yet did not reach conventional levels of significance. Adjustment for the covariates did not affect this trend (Figure 3).

**Figure 3:**
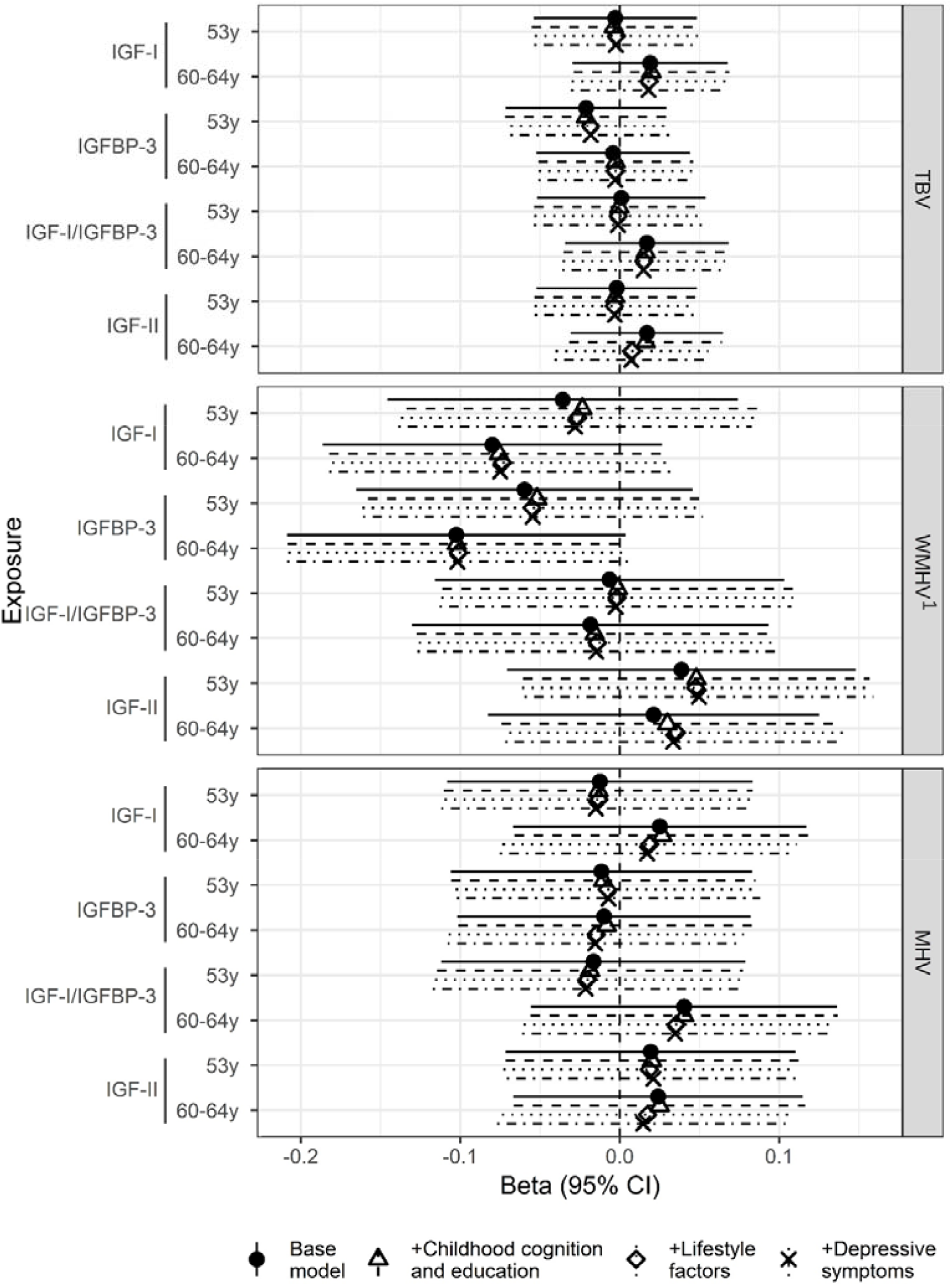
Beta coefficient and 95% confidence interval for the effect of IGF-I, IGFBP-3 and IGF-I/IGFBP-3 at ages 53y and 60-64y on Total Brain Volume (TBV), White Matter Hyperintensity volume (WMHV) and Mean Hippocampal Volume (MHV) at age 69y in the Insight 46 sub-study. Models were adjusted for sex and age at scan (base model), sex, early life cognition and lifetime education (+ childhood cognition and education model), sex, age at scan, early life cognition, lifetime education and smoking (+ lifestyle factors) and sex, age at scan, early life cognition, lifetime education, smoking and depressive symptoms (+depressive symptoms). 1Effect sizes are shown on a logarithmic scale.* significant at p < .05, ** significant at p < .01.

### IGF-I/IGFBP-3 ratio

IGF-I/IGFBP-3 ratio was associated with lower VLS and ACE-III scores (Figure 2). Elevated IGF-I/IGFBP-3 concentrations at age 60-64 were associated with lower VLS score at age 69 (ß -0.08, 95% CI -0.15,-0.02, p = 0.019) (Figure 2). In contrast to IGF-I models, adjustment for childhood cognition and education did not attenuate effect sizes. No clear associations with brain volume were observed (Figure 3).

### IGFBP-3

Higher concentrations of IGFBP-3 at age 60-64 were associated with faster processing speed at age 60-64 and 69 (ß 0.07, 95% CI 0.01,0.12, p = 0.014 and ß 0.07, 95% CI 0.02,0.12, p = 0.012, respectively) (Figure 2). The addition of covariates did not affect this association (Figure 2). Higher concentrations of IGFBP-3 at age 60-64 were additionally associated with higher scores for memory and cognitive state at age 69 (ß 0.07, 95% CI 0.01,0.12, p = 0.017 and ß 0.07, 95% CI 0.01,0.13, p = 0.033, respectively) (Figure 2). Adjustment for education and childhood cognition attenuated these estimates to ß 0.05, 95% CI 0:00,0.10, p = 0.034 and ß 0.05, 95% CI 0:00,0.10, p = 0.07, respectively (Figure 2).

Higher concentrations of IGFBP-3 at age 60-64 were additionally associated with a 10% lower WMHV at age 71 years (ß -0.1, 95% CI -0.21,0.00, p = 0.059) (Figure 3). Adjustment for the covariates did not affect this association (Figure 3).

### IGF-II

Similarly to IGF-I, higher concentrations of IGF-II were associated with better cognitive function for the WLT and ACE-III cognitive measures, but not with VLS (Figure 4). At age 53, the lowest tertile of circulating IGF-II concentration was associated with a lower ACE-III score at age 69 (score_ACE-III_ 89.48, 95% CI 88.86,90.1, p = 0.022, tertile 1) (Figure 4). Similarly, the lowest tertile of IGF-II at age 53 were associated with lower memory at age 69 (score_WLT_ 20, 95% CI 19.35,20.65, p = 0.085, tertile 1). As for IGF-I/IGFBP-3 ratio, no associations with brain volume were observed (Figure 3).

**Figure 4:**
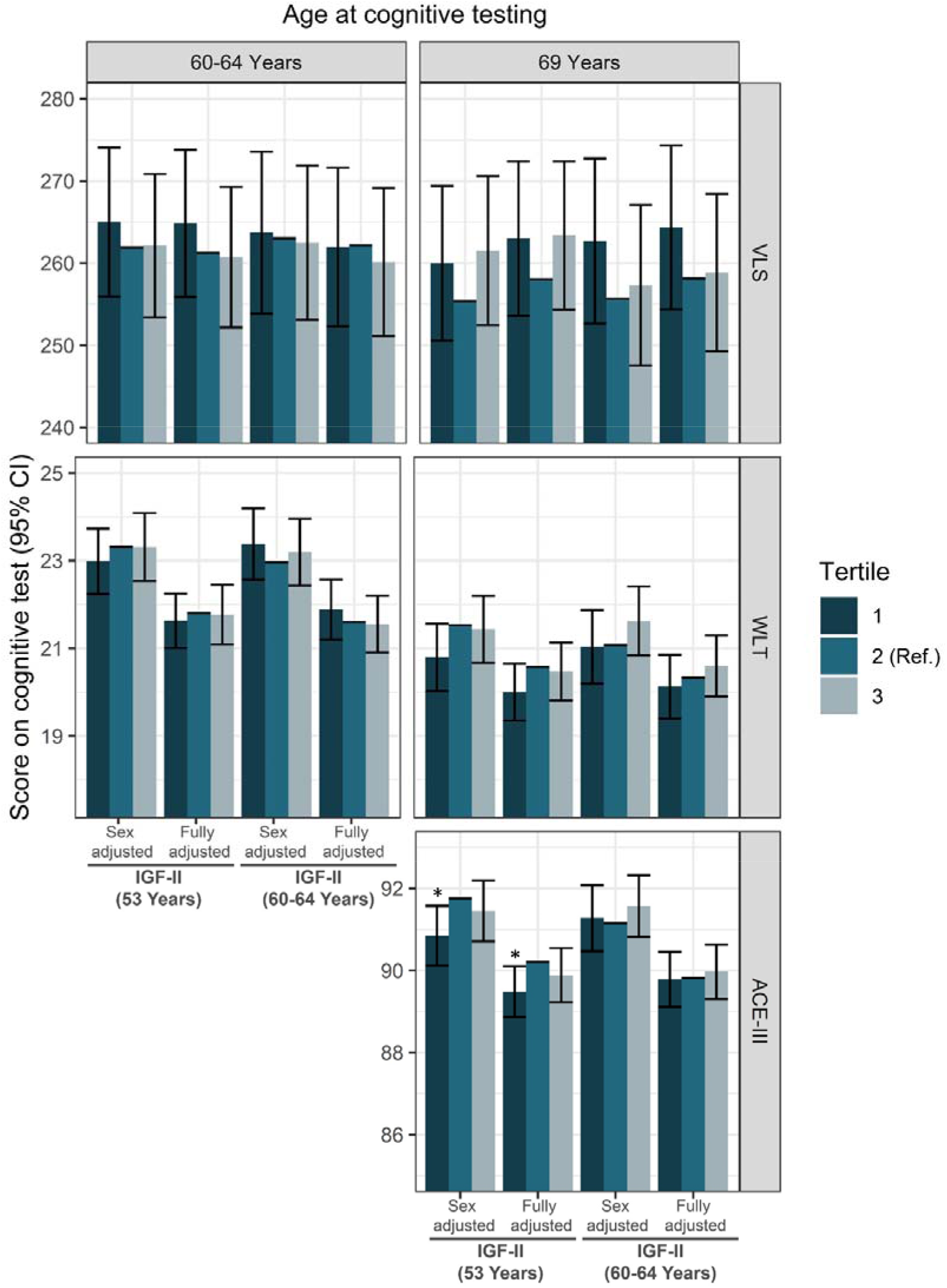
Predicted scores and 95% confidence intervals for the effect of IGF-II tertiles at ages 53y and 60-64y on ACE-III, WLT and VLS predicted score, in sex-adjusted and fully adjusted models (sex, early life cognition, lifetime education, smoking and depressive symptoms). Tertile 2 presents the reference category used in the regression. Abbreviations: Insulin-Growth Factor-I (IGF-I), Insulin-Growth Factor Binding Protein-3 (IGFBP-3), Addenbrooke’s Cognitive Exam-III (ACE-III), Word Learning Test (WLT), VLS (Visual Letter Search) test. * significant at p < .05, ** significant at p < .01.

## Discussion

In this UK population sample, IGF-I and –II concentrations at age 53, and IGF-I concentrations at age 60-64, were positively associated with cognitive state at age 69, assessed by the Addenbrooke’s Cognitive Examination (ACE-III) and verbal memory, but not with processing speed. Higher IGFBP-3 concentrations at age 60-64 were associated with overall higher cognition. Adjustment for childhood cognition and educational attainment attenuated these associations, but did not wholly account for them. In addition, IGF-I and IGFBP-3 at age 60-64 were associated with lower white matter hyperintensity burden at age 69-71. Multivariable adjustment did not notably alter these associations. Surprisingly, there was a trend towards worse cognitive state and processing speed in those with higher IGF-I/IGFBP-3 ratio levels.

The positive relationship between IGF-I and general cognitive state reflects findings in the Nurses Health Cohort (NHC), which found an association between IGF-I and verbal memory attenuated after adjustment for educational attainment ^35^. NHC participants were of similar age to NSHD participants at our 2^nd^ baseline (60-64) years, and the main follow-up point (69 years). Additionally, though, we suggest associations are similar in men and women, and show that cognitive state is related to IGF-I sampled earlier in life, at age 53. Our findings conflict with observation of a an adverse ^36^, or ‘U’ shaped relation ^37^, of IGF-I on cognition. The former analyses were performed cross-sectionally in much older participants (≥95 years), while the latter spanned ages 48-88 at follow-up. Age-dependent pleiotropic effects of IGF-I could potentially account these conflicting findings; specifically, if IGF-I acts beneficially in middle age and early older age, but has adverse effects in late older age. A ‘U’ shaped association may therefore be observed if participants span early and late older age.

The IGF pathway plays an essential role in brain development and homeostasis, including neurogenesis, neuronal differentiation and angiogenesis ^8,38^. Thus, the apparent beneficial association of increased pathway activation with cognitive and brain outcomes may be driven by a combination of developmental – with better cerebral development during foetal and early life and neuroprotective pathways – with increased neurogenesis and improved outcomes following cerebrovascular events ^8,39–41^. Prospective analyses of the Framingham Heart Study found that higher concentrations of IGF-I were associated with larger brain volumes in healthy participants, potentially suggestive of some protection against neurodegeneration ^42^. Association magnitudes were doubled in the parent versus the offspring cohort (mean ages 79 years and 61 years, respectively), highlighting an important age dependency. Whilst we did not observe an association between IGF-I and TBV, IGF-I was associated with lower WMHV, an indicator of cerebral small vessel damage. To our knowledge, no previous study has explored the association between IGF-I and cerebral small vessel disease. A 10-year prospective study found stroke incidence to be associated with lower circulating IGF-I concentrations ^43^. Other studies have shown improved outcomes following ischemic stroke in participants with higher IGF-I concentrations (41). In hypertensive mice, liver-specific IGF-I-knockdown treatment was associated with increased and earlier incidence of cerebral microhaemorrhages compared to control mice, likely due to impaired remodelling processes ^45^. Mechanisms underlying protective effects of IGF-I on the vasculature include anti-inflammatory properties, preservation of endothelial function, and a positive association with cardiovascular risk factors such as obesity and insulin resistance ^43^. Whilst our findings did not reach conventional levels of statistical significance, effect sizes, consistency with previous research, and strong biological plausibility supports further investigation.

Surprisingly, however, associations with the IGF-I/IGFBP-3 ratio were opposite to those observed for IGF-I alone, with the ratio associated with reduced processing speed and, to a lesser degree, cognitive state. This can be explained by IGFBP-3 being positively associated with cognition independently of IGF-I. IGFBPs have been shown to execute IGF-I independent actions, including mediation of cell proliferation, apoptosis and survival ^46,47^. Furthermore, the age-related decline in IGFBP-3 concentration was markedly greater than that for IGF-I resulting in an increase in IGF-I/IGFBP-3 ratio between the two timepoints. Thus, the apparent negative association with cognitive outcomes may be the product of the differential declines in the ratio components rather than a biologically relevant association of the ratio with the cognitive outcomes.

We observed globally improved cognitive outcomes associated with increased circulating IGFBP-3 concentrations. In line with our results, higher concentrations of IGFBP-3 were found to be cross-sectionally associated with better cognitive state in women in the Mayo Clinic Study of Ageing and reduced dementia incidence in men ^48,49^. In mice, null mutations in the *Igfbp3* gene resulted in impaired neuronal structure and signalling and spatial working memory ^50^. However, findings are not consistent across studies with higher concentrations of IGFBP-3 related to poorer cognition in males in the Caerphilly Prospective Study (CaPS) ^10^. Our findings highlight the importance of exploring associations with individual factors that form a biologically meaningful molecular ratios, and understanding age-related changes in exposures, which, as we show, may not be equivalent across associated measures.

In addition to favourable cognitive outcomes, IGFBP-3 was conjointly associated with a 10% reduction in WMHV in our study. Several studies have implied the possibility of a beneficial association between IGFBP-3 and cerebral small vessel disease. High concentrations of circulating IGFBP-3 are associated with reduced risk of ischaemic stoke and improved functional outcomes following stroke events ^44,51,52^. In mice, IGFBP-3 was shown to protect retinal vasculature from hyperoxia-induced vessel regression and, *in vitro*, promoted differentiation of endothelial precursor cells to endothelial cells, indicating a potential pathway by which IGFBP-3 may reduce WMHV ^5,53^.

Few studies have examined the relationship between IGF-II and cognitive outcomes. IGF-II has traditionally been seen as relatively unimportant beyond fetal development. However, there is increasing evidence relating detrimental IGF-II expression to disease after birth ^4,54^. To our knowledge, only one other study has investigated the role of IGF-II on later-life cognition. The CaPS found a reduced occurrence of cognitive impairment without dementia with higher circulating IGF-II concentrations, but not with cognitive state. In addition, they did not find an association between IGF-I and cognitive function ^10^. In our study, we found a time-dependent association between higher IGF-II at age 53 and better visual memory and cognitive state at age 69, presenting similarities to the results obtained from the IGF-I models. This suggests a common pathway, likely via signalling downstream of the IGF-1R. Discrepancies between our findings and those from the CaPS may be driven by choice of outcome. The CaPS studied CIND, dementia and ‘normal’ cognitive ageing separately, therefore excluding participants with CIND or dementia from models studying ‘normal’ cognitive ageing. It is possible that ‘normal’ and ‘abnormal’ cognitive ageing are driven by separate mechanisms and IGF-I and IGF-II might be protective in the context of ‘abnormal’ cognitive ageing only.

Our study has some important strengths, including a prospective design with extended follow-up (16 years) and important confounders, including childhood cognition. To our knowledge, it is the only study to use repeat exposure and outcome measures to better understand the temporal relationship between IGFs and cognition. Age-dependent relationships may reflect separate underlying biological pathways acting at different points in the life course. In addition, it is the first study to investigate both cognitive function and structural brain measures. However, it is important to consider some limitations of the present study. Whilst associations were robust to adjustments, the observational methods used here cannot discount the possibility of residual confounding. For instance, this study did not adjust for chronic diseases which may impact IGF serum levels and associate with brain health independently of the IGF pathway ^55^.

To investigate active IGF-I, this study used the ratio of IGF-I to IGFBP-3. However, we acknowledge this to be an imprecise measure of IGF-I activity, as other binding proteins and pathway components are involved in regulating pathway activation. Furthermore, binding proteins have been shown to have IGF-I independent actions46. Conflicting results between IGF-I and IGF-I/IGFBP-3 in our study were seemingly explained by IGF-I-independent associations between IGFBP-3 and cognition, thereby supporting future study of IGFBP-3 independently from IGF-I. However, the use of ‘free’ IGF-I may provide a more accurate view of the role of unbound circulating IGF-I on brain health ^56^.

Blood samples from ages 53 and 60-64 were assayed together and stored at -80°C until quantification. Therefore, measures will have been stored for varying lengths of time which may affect results, particularly if the protein is prone to degradation. However, IGF-I, -II and IGFBP-3 levels have previously been shown to remain stable following 9 years of -80°C storage, which reduced the likelihood of this occurring in our study ^57^. Furthermore, biomarker concentrations may be affected by the differential fasting status when blood samples were collected at ages 53 (non-fasting) and 60-64y (fasting). However, Bereket *et al* (1996) found no associations between IGF-I and IGFBP-3 concentration and short-term fasting thus limiting the risk of fasting status being an important biasing factor in our study ^58^. Finally, with the latest follow-up at age 69, participants are still not old enough for severe cognitive decline to be common, which could mean limited statistical power to detect moderate to small associations.

In light of these findings, we would suggest further studies to determine the exact domains which may be affected by IGF-I, –II and IGFBP-3 concentrations. This could be done by using an array of cognitive tests that probe distinct cognitive domains. Further studies continuing to use repeat IGF and IGFBP exposures in relation to cognition measures across the life course may help provide a better understanding of the complex relationship between the two. Re-examination of the relationships in the NSHD at an older age would be essential to understand age-dependent effects and potentially address the current inconsistencies in the field. Furthermore, genetic approaches to study the association between polygenic risk scores and cognition across the age spectrum may provide an alternate method to investigate relationships across the life course.

While repeat measures in our study have highlighted the importance of time of exposure, outcome and follow-up period, further work is required to disentangle the temporal nature of the IIS-brain health relationship. Regular IGF-I, -II, IGFBP-3 and cognition measures throughout the life course would help to elucidate this. As autocrine and paracrine IGF-I concentrations vary with age, narrowing the age range at each sample collection could also prove essential ^59,60^. The wide age range in a number of studies may prove responsible for some of the heterogeneity currently observed in results ^37,56,61^.

In conclusion, in this UK birth cohort, higher circulating IGF-I and -II levels were prospectively associated with a favourable cognitive state, whilst IGF-I/IGFBP-3 ratio was associated with slower processing speed. IGF-II was additionally associated with better memory performance. IGFBP-3 was independently associated with cognitive state, memory, processing speed and WMHV. The present study has not only added to our understanding of the IIS-cognition relationship, but crucially, it has elucidated the importance of using a breadth of neuroimaging, cognitive function and state to understand more holistically how IGFs and IGFBPs relate to brain health from mid to later life.

## Data Availability

Restrictions apply to the availability of some or all data generated or analyzed during this study to preserve patient confidentiality or because they were used under license. The corresponding author will on request detail the restrictions and any conditions under which access to some data may be provided.

## Acknowledgements

The authors are grateful for the continued support of the NSHD study members without whom this study would not have been possible. We would also like to acknowledge Dr. Andy Wong for assistance in data extraction and management, and the Insight 46 study team.

